# Reconstructing the phylodynamic history and geographic spread of the CRF01_AE-predominant HIV-1 epidemic in the Philippines from PR/RT sequences sampled from 2008-2018

**DOI:** 10.1101/2023.01.04.22283982

**Authors:** Francisco Gerardo M. Polotan, Carl Raymund P. Salazar, Hannah Leah E. Morito, Miguel Francisco B. Abulencia, Roslind Anne R. Pantoni, Edelwisa S. Mercado, Stéphane Hué, Rossana A. Ditangco

**Author notes:** SH and RD contributed equally.

## Abstract

The Philippines has had a rapidly growing HIV epidemic with a shift in the prevalent subtype from B to CRF01_AE. However, the phylodynamic history of CRF01_AE in the Philippines has yet to be reconstructed. We conducted a descriptive retrospective study reconstructing the history of HIV-1 CRF01_AE transmissions in the Philippines through molecular epidemiology. Partial polymerase sequences (*n* = 1429) collected between 2008 and 2018 from 13 Philippine regions were collated from the RITM drug resistance genotyping database. Subsampling was performed on these Philippine and Los Alamos National Laboratory HIV international sequences followed by estimation of the time to the most recent common ancestor (tMRCA), effective reproductive number (*R_e_*), effective viral population size (*N_e_*), relative migration rates and geographic spread of CRF01_AE with BEAST. *R_e_* and *N_e_* were compared between CRF01_AE and B. Most CRF01_AE sequences formed a single clade with a tMRCA of 1999 [95% HPD: 1996, 2001]. Exponential growth of *N_e_* was observed from 1999 to 2013. The *R_e_* reached peaks of 3.71 [95% HPD: 1.71, 6.14] in 2009 and 2.87 [95% HPD: 1.78, 4.22] between 2012 and 2015. A transient decrease to 0.398 [95% HPD: 0.0105, 2.28] occurred between 2010 and 2012. The epidemic most likely started in Luzon in the National Capital Region, which then spread diffusely to the rest of the country. Both CRF01_AE and subtype B exhibited similar but unsynchronized patterns of *R_e_* over time. These results characterize the subtype-specific phylodynamic history of the CRF01_AE epidemic in the Philippines, which contextualizes and may inform past, present, and future public health measures toward controlling the HIV epidemic in the Philippines.

## Introduction

Human immunodeficiency virus-1 (HIV-1) infections and deaths related to acquired immunodeficiency syndrome (AIDS) have been rapidly increasing in the Philippines over the past years, with a 237% percent change in new infections, the highest in Asia and the Pacific from 2010 to 2020 [1]. As of June 2022, 101,768 confirmed HIV cases have been reported since January 1984, with about 92% of these reported in the last 10 years and the number of new diagnoses increasing from 6 cases/day in 2011 to 41 cases/day in 2022 [2].

The demographics of HIV-1 cases in the Philippines have also shifted over time. From 1984 to 1990, the majority of diagnosed cases (62%) were females, compared to a majority of male cases (94%) in 1991–2020 [3]. The largest proportion of new cases also shifted to a younger age group, from 35–49 in 2001–2005 to 25–34 and 15–24 age groups in 2010–2022 [2], [4], [5], [6]. Furthermore, the majority of transmissions were from male-to-female sex until 2009, pointing to sexually active men who have sex with men (MSM) as the major transmitters of HIV-1 in the Philippines since 2010 [5].

The composition of circulating subtypes in the Philippines has also changed. In 1998, the major subtype was B, with around 70% of infections, followed by the CRF01_AE, a putative recombinant of subtypes A and E, with 16–29% of infections [7], [8]. However, the major subtype has shifted to CRF01_AE, making up 77% of HIV-1 patients in a 2013 cohort and with 22% of the same cohort being subtype B [8]. A more recent cohort from 2016 to 2018 reported the distribution of 77% CRF01_AE, 13.8% subtype B, and 9.2% other subtypes or recombinants [9]. Furthermore, CRF01_AE was the major subtype among the MSM population, while B was predominant among injecting drug users (IDUs) in infections from 2007 to 2012 [10], [11]. It is important to highlight that the HIV epidemic in the Philippines is predominantly CRF01_AE, which displays high antiretroviral resistance [12], and thus treatment regimens may need readjustment since recommendations are based primarily on clinical trials from regions with subtype B infections [13], [14].

CRF01_AE may have been introduced into the Philippines around the mid-1990s, potentially from a neighboring Asian or Southeast Asian country [15], [16]. Despite the predominance of CRF01_AE among the circulating subtypes in the Philippines, to the best of our knowledge, there has not yet been any study disentangling epidemiological parameters of specific HIV subtypes in the Philippines using phylodynamic methods.

Although whole genome sequences would be able to more accurately distinguish recombinant sequences [17] and lead to more precise estimates [18], partial *pol* sequences from drug resistance genotyping (DRG) have been shown sufficient to reconstruct transmission histories phylogenetically and are highly available data from routine testing [19]. The Research Institute for Tropical Medicine (RITM) DRG database has archived *pol* protease/reverse transcriptase (PR/RT) sequences from routine testing for over a decade from referred samples spanning multiple Philippine regions, compiling a suitable and available dataset for reconstructing subtype-specific phylodynamics.

Thus, we aimed to reconstruct CRF01_AE spatiotemporal dynamics using phylogenetic analysis of available partial *pol* sequences consisting of the PR/RT regions collected from patient samples referred to the RITM, one of the largest referral hubs in the Philippines, between 2008 and 2018. We investigated the dates of introduction events; historical changes in epidemiological parameters of effective viral population size, effective reproductive number, geographical spread, and migration rates of CRF01_AE; and how these compared with the *N_e_* and *R_e_* of subtype B, to distinguish the relative contribution each subtype has had to the overall HIV epidemic and provide important context to public health policies attempting to control its spread in the country.

## Methods

### Study population and sample selection

The retrospective study population were HIV-positive cases from the in-house RITM HIV-1 DRG database with 1530 cases matching the inclusion criteria (see the following section) for CRF01_AE analysis, while 265 cases matched the inclusion criteria for the comparative subtype B analysis. The data included PR/RT Sanger sequences generated from RITM routine DRG with accompanying information on specimen collection date. Collated data for CRF01_AE analysis consisted of sequences from patient samples from RITM and from referring hospitals and social hygiene clinics from 13 out of 17 administrative regions from all three Philippine island groups between January 2008 and November 2018. Meanwhile, data for the subtype B analysis came from RITM patients and patients from referring hospitals and social hygiene clinics from 10 out of 17 administrative regions from all three Philippine island groups between February 2014 and February 2020.

Demographic data (i.e., sample collection date and requisitioner address as location) were obtained via request forms. Missing demographic data from the samples were requested from the Department of Health Epidemiology Bureau. People living with HIV have unique HIV laboratory test ID numbers, and these were used to obtain data from the Epidemiology Bureau without using patient names.

The Institutional Review Board (IRB) of the Research Institute of Tropical Medicine waived ethical approval for this study by granting the study protocol a certificate of exemption from review as it was a retrospective analysis of archived de-identified routine DRG sequence data and metadata only.

### Subtype classification and sequence alignment

The inclusion criteria used were: all PR/RT *pol* sequences available in the RITM MBL DRG; sequences from RITM classified as CRF01_AE or subtype B; sequences at least 500 nucleotides long. The exclusion criteria were: non-CRF01_AE or non-subtype B sequences from RITM MBL DRG, sequences less than 500 nucleotides long; Los Alamos National Laboratory (LANL) HIV Sequence Database [20] sequences without country and year of sampling information; sequences containing ≥5% ambiguous nucleotides, frameshift mutations, stop codons, and APOBEC-mediated hypermutations.

Thus, partial *pol* sequences from the RITM DRG database (*n* = 1621) were classified according to HIV subtype (i.e. phylogenetically related strains defined by close genetic distance with each other) using the Stanford HIV Drug Resistance Database [21] and HIVdb program (v8.6) [22], and programs COMET [23] and REGA [24] were used to exclude non-CRF01_AE and recombinant sequences, respectively. The 10 most closely related CRF01_AE *pol* gene sequences (HXB2 coordinates 2253-3233) per input sequence were also retrieved from the LANL HIV Sequence Database [20] in January 2019 using HIV-BLAST. LANL sequences (*n* = 454) from 1990 to 2016 with information on the sampling year and country and without >5% ambiguous nucleotides, frameshift mutations, stop codons, and APOBEC-mediated hypermutations were retrieved. Sequences that contained frameshift insertions/deletions as determined by HIVdb were excluded. A small number of sequences that had very few bases in the PR/RT region or had very long insertions were also excluded. A codon alignment of all remaining sequences was produced with the Gene Cutter tool from LANL [25]. A 129 bp-long stretch of sequence, spanning reference HXB2 codon position 96 of the protease gene to codon position 39 of the reverse transcriptase gene, was removed from the alignment since a majority of the Philippine sequences in this study contained gaps in this region. Drug resistance-associated codons were stripped to remove the influence of convergent evolution from drug resistance mutations [26]. Large overhangs were trimmed using Aliview [27], and sequences from the same patient were removed, resulting in 1429 sequences.

Using similar procedures, high-confidence subtype B sequences were identified (*n* = 265) for the comparative analysis. The 10 most similar subtype B *pol* sequences per input sequence were also retrieved from LANL in May 2021 using HIV-BLAST (*n* = 599). Codon alignment, filtering, and trimming were also performed. Five partial polymerase sequences each from of subtype A (DQ396400, DQ445119, JQ403028, KX389622, and MH705133) and subtype C (AB023804, AB097871, AB254143, AB254146, and AB254150) were included as outgroups, leading to 874 sequences.

For both CRF01_AE and subtype B sequences, collection dates with complete year and month but not day were imputed to the first day of the month, while collection dates with complete year only but not month and day were imputed to the first month and day of the year.

### Phylodynamic analysis

FastTree2 [28] and IQ-TREE2 [29] were used to reconstruct a maximum likelihood phylogeny from the full CRF01_AE alignment, and TempEst2 [30] was used to measure the temporal signal of the sequence alignment by root-to-tip regression analysis. A manual subsample of sequences from the largest Philippine clade and international sequences nearest to the clade (*n* = 200) was used to estimate the time of introduction of CRF01_AE into the Philippines based on the time to the most recent common ancestor (tMRCA) of that clade. Only 200 sequences were selected to reduce computation time. BEAST v2.6.7 [31] was used to estimate the tMRCA of the CRF01_AE clade, with a Coalescent Bayesian Skyline Plot (BSP) tree prior [32], bModelTest [33] set as the nucleotide substitution model, clock rate prior set to a normal distribution (M = 1.5E-3, S = 4.9E-4) [34]–[37], clock rate standard deviation prior to an exponential distribution (M = 0.1), and the best-fitting molecular clock model of either strict or relaxed clock [38] determined by the path sampling and stepping-stone procedure [39], [40] implemented in BEAST v1.10.4 [41]. Five separate Markov chain Monte Carlo (MCMC) chains were run for 10M states each, sampling every 1000 states. These were combined and downsampled with LogCombiner [42] to 10,000 states and trees. TreeAnnotator [42] was used with a 10% burnin to generate the maximum clade credibility (MCC) tree.

To estimate the effective viral population size (*N_e_*) and effective reproductive number (*R_e_*) of CRF01_AE across time, the subset of Philippine sequences belonging to the large Philippine monophyletic clade was used. Sequences with missing sample collection dates or location data were excluded from the analysis. Sequences from this clade were subsampled uniformly (Supplemental File XLSX) across time and geographic location [43], [44] according to island group (i.e., Luzon, Visayas, and Mindanao) and between May 2008 and November 2018. Specifically, the subsampling procedure outlined by Hidano et al was used [44], producing a subset of 260 sequences. BEAST2 was run with the BSP and Birth-Death Skyline (BDSKY) Serial [45] tree priors, MCMC chain lengths of 400M and 300M states, sampling every 40,000 and 30,000 states to estimate *N_e_* and *R_e_*, respectively. bModelTest [33] was selected as the nucleotide substitution model, the clock rate prior set to as indicated previously, and the best-fitting molecular clock model determined by the path sampling and stepping-stone procedure. For the BSP analysis, the bPopSizes and bGroupSizes were both set to the dimension of 5. For the BDSKY analysis, the origin of the epidemic prior was set to a log normal distribution (M=3.4, S=0.29), the become uninfectious rate prior to a log normal distribution (M = 2.08, S = 1.0), the sampling proportion prior to a normal distribution (mean = 0.004, sigma = 0.001), the reproductive number prior to a log normal distribution (M = 0.0 in log space, S = 1.25), and the reproductive number dimensions to 10. The MCC tree was generated from trees sampled in the BDSKY analysis using TreeAnnotator [42] with 10% burnin. The bdskyTools R package was used to smooth the *R_e_* estimates over time [46].

To compare the CRF01_AE *N_e_* and *R_e_* with those of another circulating subtype that may reflect similar patterns if non-subtype-specific influences were acting upon their transmission, an equivalent analysis was done using BEAST2 on the largest subtype B monophyletic clade identified from a phylogeny reconstructed using IQ-TREE 2. No subsampling was performed as only 195 sequences sampled from September 2008 to February 2020 were available in this clade. The same respective priors as CRF01_AE were used, with bModelTest [33] selected as a nucleotide substitution model and with the best-fitting molecular clock model also determined by the path sampling and stepping-stone procedure.

### Phylogeographic analysis

We performed a Slatkin–Maddison test [47] on the FastTree2 phylogeny of all sequences from the large monophyletic Philippine CRF01_AE clade to test for significant associations (α = 0.05) between the trait of location (island group and region) and tree topology. We also tested for significant associations (α = 0.05) of island group and tree topology using the Bayesian Tip-Significance Testing (BaTS) package [48], which calculates the Association Index (AI), Parsimony Score (PS), and Monophyletic Clade (MC) index statistics, on 1000 downsampled trees from the BSP analysis of uniformly subsampled CRF01_AE sequences and 999 null replicates. BEAST v1.10.4 [41] was used for the phylogeographic analysis of the CRF01_AE alignment with nucleotide positions and demes set as the first and second partitions, respectively. Analysis was performed using a strict clock model and a BSP tree prior. The asymmetric discrete trait substitution model with BSSVS was selected for using Philippine island groups as demes. The symmetric discrete trait substitution model with BSSVS was selected for using these island groups from the same subsampled data converted to Philippine regions (NCR, I, II, III, IV-A, V, VI, VII, IX, X, XI, XII, and XIII) as demes. SpreaD3 [49] was used to visualize phylogeographic migration at eight different time points and at both island group and regional levels over a geoJSON map of the Philippines [50].

All other settings besides those stated were left at default values. See complete details of the models and parameters used for each analysis in the Supplemental File XLSX. An effective sample size (ESS) of ≥200 was deemed as satisfactory convergence of the estimated parameters. The TRACER [51], FigTree [52], and IcyTree [53] software and R packages “ggtree” [54] and “ggplot2” [55] were used to visualize the results.

## Results

### Introduction of CRF01_AE to the Philippines in the late 1990s to early 2000s

The majority CRF01_AE sequences sampled in the Philippines (1150/1185; 97%) belonged to one large monophyletic clade (Fig. 1a; SH-aLRT, UFBoot branch support: 98.3, 100). Fewer sequences (35/1185; 3%) were singletons or belonged to smaller clusters of at most three sequences, suggesting multiple introductions from overseas over time. Furthermore, the large monophyletic Philippine CRF01_AE clade contained sequences from the United States, Japan, Hong Kong, Australia, Great Britain, China, and South Korea, suggesting transmission to and/or from these countries either directly or involving unsampled intermediate destinations (Fig. 1a). Root-to-tip linear regression on the full set of sequences showed a positive correlation between sequence diversity and time of sampling (coefficient of correlation: 0.74; *R*^2^ = 0.55), indicating sufficient temporal signal for further molecular clock estimations (Fig. 1b). Model selection through path sampling and stepping-stone procedure on a subsample of 200 sequences indicated greater support for a relaxed molecular clock over a strict clock. Under this model, the large Philippine cluster from the subsampled alignment had a mean tMRCA of March 1999 [95% HPD: April 1996, December] (Fig. 1c). Additionally, the estimated evolutionary rate for this international set of sequences was a mean of 2.413E-3 [95% HPD: 2.0457E-3, 2.7851E-3] nucleotide substitutions/site/year.

**Figure 1.**
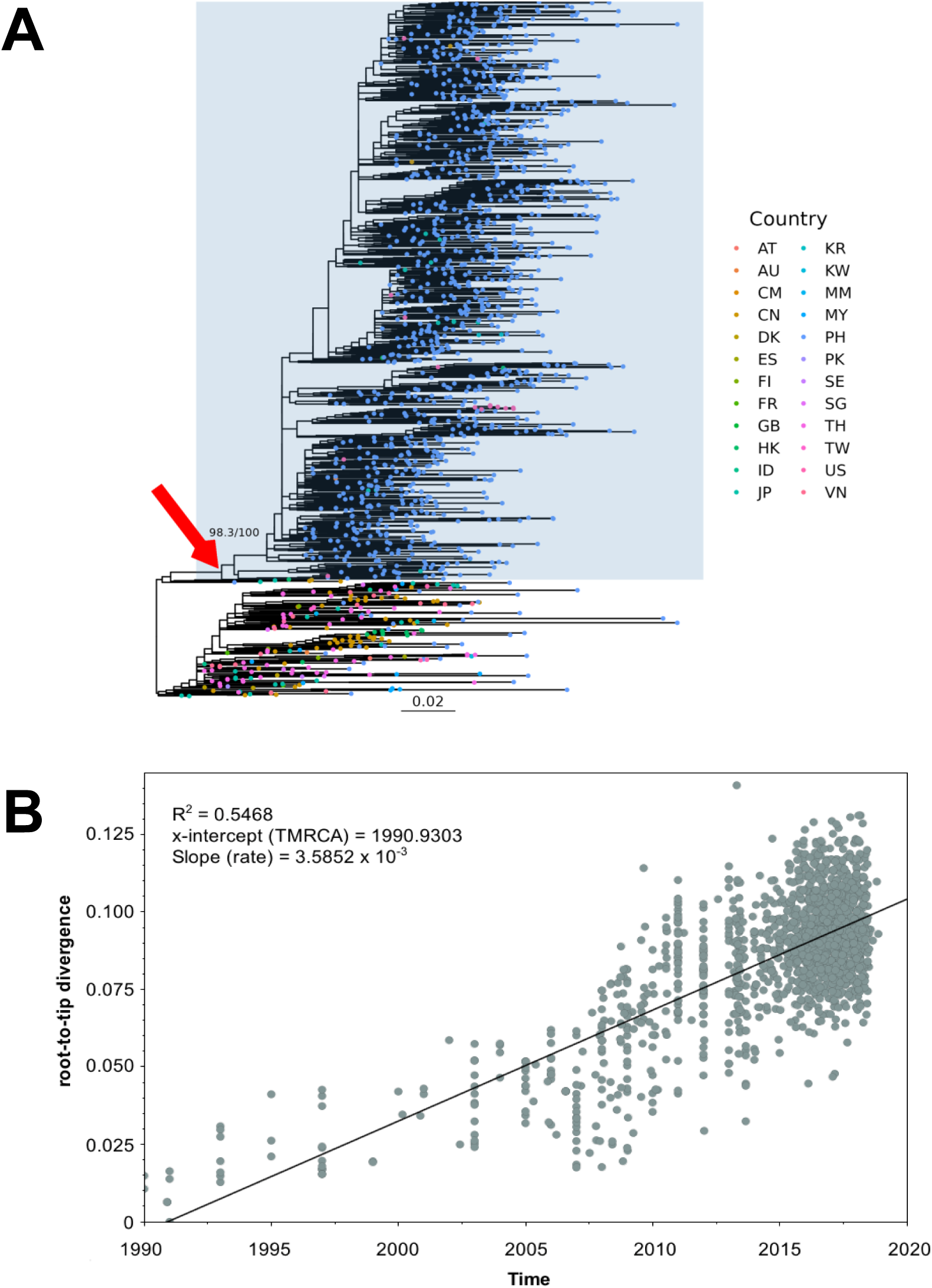
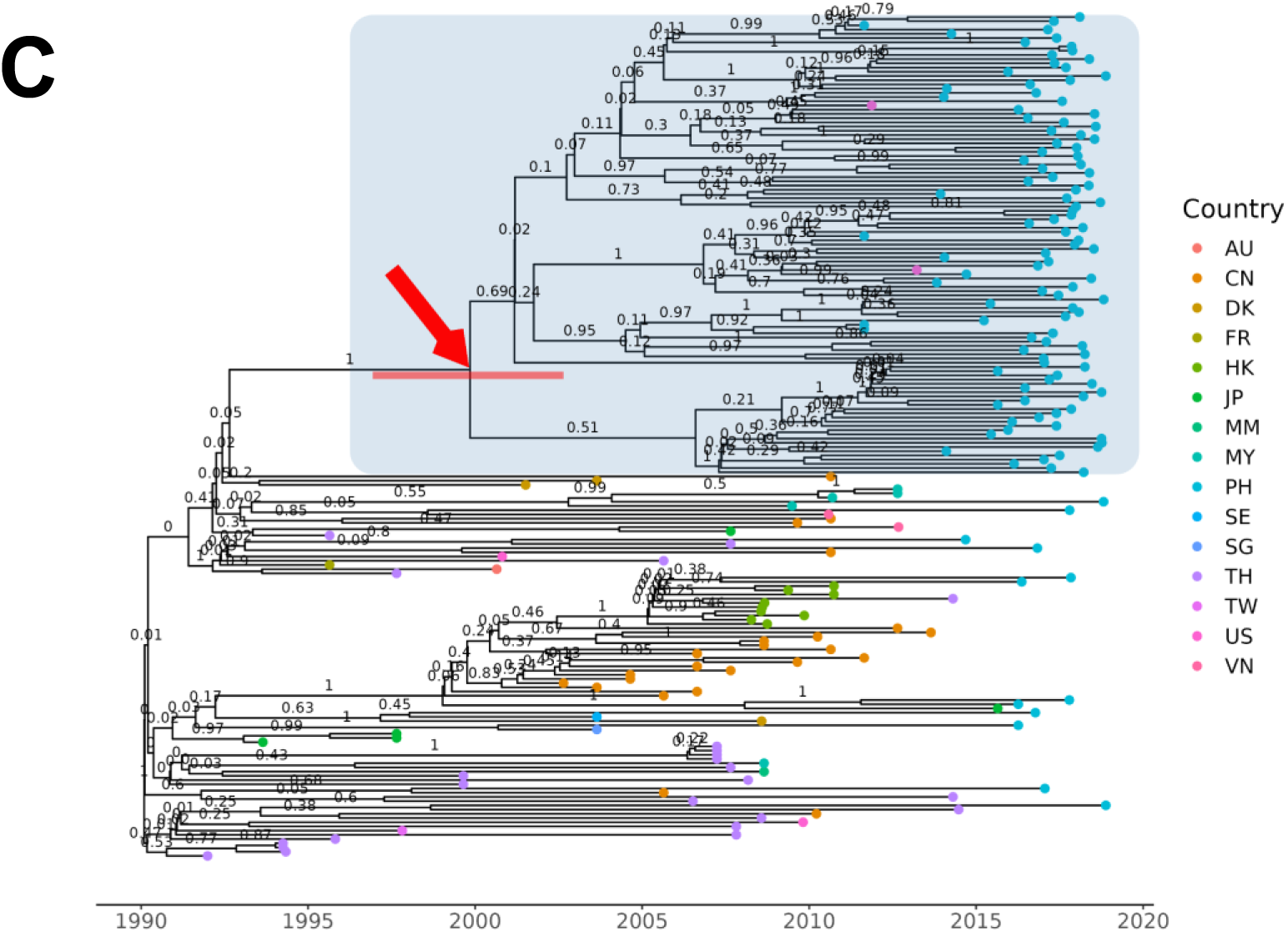
(A) Maximum likelihood phylogenetic tree of Philippine CRF01_AE PR/RT sequences relative to international sequences obtained from the LANL database reconstructed with IQ-TREE2. The red arrow indicates the node, with SH-aLRT and UFBoot branch support values, of the most recent common ancestor of the large monophyletic clade to which the majority of CRF01_AE sequences belong. This clade is also emphasized with a blue rectangular highlight. The bottom scale bar shows a reference branch length for 0.02 substitutions/site. Country abbreviation: AT:Austria; AU:Australia; CM:Cameroon; CN:China; DK:Denmark; ES:Spain; FI:Finland; FR:France; GB:United Kingdom; HK:Hong Kong; ID:Indonesia; JP:Japan; KR:South Korea; KW:Kuwait; MM:Myanmar; MY:Malaysia; PH:Philippines; PK:Pakistan; SE:Sweden; SG:Singapore; TH:Thailand; TW:Taiwan; US:United States; VN:Vietnam. (B) Root-to-tip plot generated with TempEst showing a positive correlation between time and divergence or accumulating mutations among sequences, indicating suitability of sequence data for time-scaled phylogenetic and phylodynamic analysis. (C) Time-scaled phylogenetic tree generated using BEAST2 from a manual subset of the monophyletic Philippine CRF01_AE sequences and contextual international LANL sequences. The red arrow indicates the node of the most likely time to the most recent common ancestor for the Philippine CRF01_AE sequences, along with an error bar for the uncertainty in estimated time. This clade is also emphasized with a blue rectangular highlight.

### The growth of CRF01_AE in the Philippines

Coalescent BSP analysis was conducted with 260 subsampled sequences from the large Philippine monophyletic clade. Model selection with path sampling and stepping-stone procedure indicated greater support for a strict clock over a relaxed clock (Supplemental File XLSX). The resulting Skyline plot, showing changes of effective population size over time, revealed an exponential growth phase that lasted about 14 years between 1999 and 2013 whereby *N_e_* increased by four orders of magnitude (Fig. 2b). The peak of this growth phase was followed by a stable or plateau phase wherein *N_e_* remained within the same order of magnitude from 2013 onwards (Fig. 2b).

**Figure 2.**
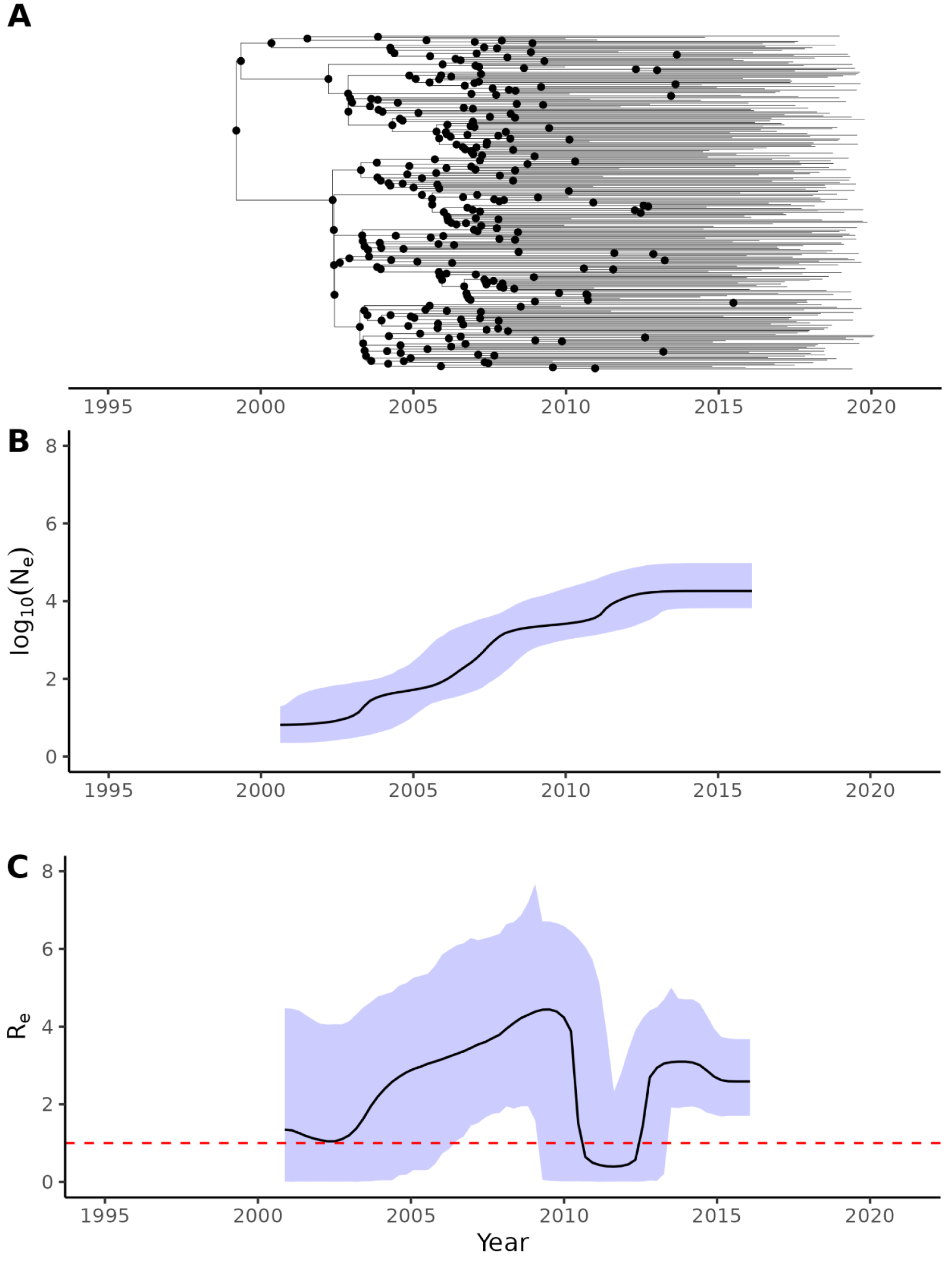
Phylodynamics of HIV CRF01_AE in the Philippines measured between mid-1990s and 2018 reconstructed by BEAST2 analysis of uniformly sampled Philippine CRF01_AE PR/RT sequences. (A) Time-scaled maximum clade credibility tree from BDSKY analysis summarized with TreeAnnotator. Common ancestor nodes are highlighted as black dots. (B) The change in CRF01_AE effective population size (*N_e_*) over time in log scale, with the median represented as a black line and the 95% HPD as a blue-shaded interval, obtained from BSP analysis. (C) The change in the effective reproductive number (*R_e_*) over time, with the mean represented as a black line and the 95% HPD as a blue-shaded interval, obtained from BDSKY analysis. A dashed red line indicates the value of *R_e_* equal to 1.

### Fluctuations of CRF01_AE reproductive number from the late 1990s to 2016

Phylodynamic analysis was performed with the same 260 subsampled CRF01_AE sequences and the best-fitting strict clock model. Between the late 1990s and 2016, the confidence intervals of the estimated *R_e_* of CRF01_AE in the Philippines spanned 1.0 for two periods, wherein the epidemic or the number of secondary cases per infected case remained stable, and was significantly above 1.0 for two periods, during which there was exponential growth of the epidemic (Fig. 2b). In more detail, *R_e_* increased from about 1.06 [95% HPD: 0.0210, 3.86] to a peak of about 3.71 [95% HPD: 1.71, 6.14] from February of 2000 to April of 2008, then decreased between 2009 and 2013 to as low as 0.398 [95% HPD: 0.0105, 2.28], and finally rebounded up to about 2.87 [95% HPD: 1.78, 4.22] in December of 2013, where it remained until 2016, the end of the interval with informative common ancestor nodes from the dataset (Fig. 2b).

### Transmission of CRF01_AE from the National Capital Region to other Philippine island groups and regions

The Slatkin–Maddison test performed on all Philippine sequences from the large CRF01_AE transmission cluster detected significant clustering of sequences by island group (96 observed transitions, 109–117 min–max null model transitions; *p*-value <0.001) and by Philippine region (152 observed transitions, 163–171 min–max null model transitions; *p*-value <0.001) (Fig. S1). Similarly, with the BaTS test, significant clustering by island group was obtained for the AI (7.82-10.45 observed 95% CI, 12.86–15.08 null 95% CI, *p*-value <0.001), PS (50.00–58.00 observed 95% CI, 69.33–74.42 null 95% CI, *p*-value <0.001), MC Luzon (11.00–17.00 observed 95% CI, 6.44–10.29 null 95% CI, *p*-value = 0.014), MC Visayas (2.00–3.00 observed 95% CI, 1.21–2.08 null 95%, *p*-value = 0.002), and MC Mindanao (4.00–6.00 observed 95% CI, 1.96–3.05 null 95% CI, *p*-value = 0.0010) statistics (Table S2).

Phylogeographic analysis was performed with the same 260 subsampled CRF01_AE sequences and the best-fitting strict clock model. The most likely location of the root of the CRF01_AE phylogeny was estimated to be either the Luzon island group (posterior probability = 1.0) (Fig. 3a) or the NCR administrative division (posterior probability = 0.9992) (Fig. S2a), implicating these locations as the origin of the CRF01_AE epidemic. No significant differences were observed among the estimated relative migration rates between the three different island groups (Fig. 3b).

**Figure 3.**
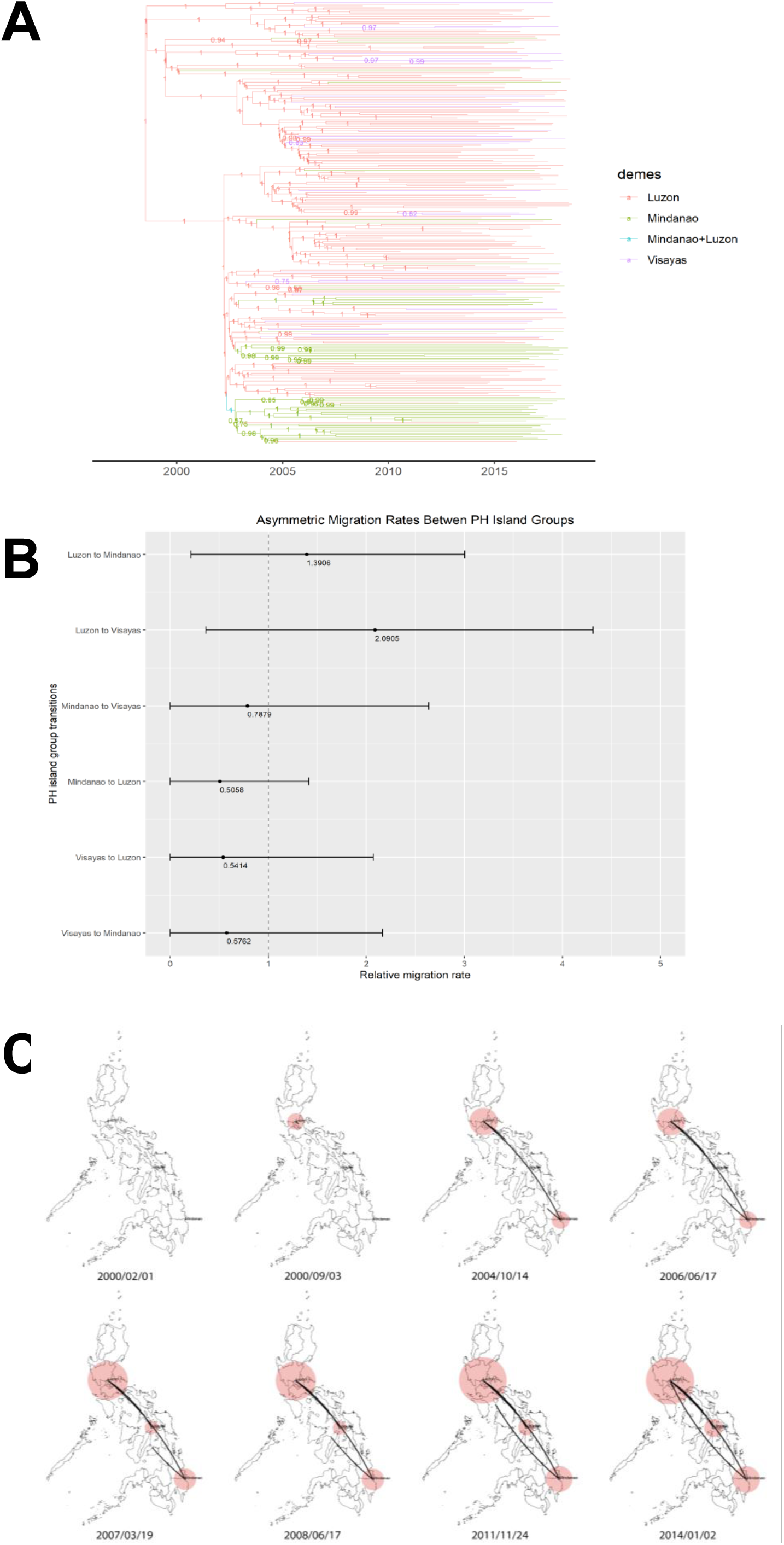
Analysis of geographic spread and relative migration rates of HIV CRF01_AE across Philippine island groups. (A) Maximum clade credibility tree of monophyletic Philippine CRF01_AE PR/RT sequences generated with BEAST under the “phylogeo” model and summarized with TreeAnnotator. Branches are labeled with posterior probability support values of corresponding nodes and are colored according to the most likely geographic location of a branch at the level of Philippines island groups: Luzon (red), Visayas (green), Mindanao (purple), and Mindanao+Luzon (blue). (B) Forest plot with mean and 95% HPD estimate of the asymmetric relative migration rates of CRF01_AE between all pairs of Philippine island groups. A dashed line indicates a relative migration rate equal to 1.0, or no greater or lesser than other migration rates. (C) Phylogeographic spread of CRF01_AE between Luzon, Visayas, and Mindanao island groups over time visualized with SPREAD3. The size of the red polygons over island groups correspond to the intensity of localized virus transmission at the specified location and time.

The reconstructed phylogeographical spread of CRF01_AE over time indicated low local spread in NCR (Luzon) around early 2000 during the estimated start of the epidemic followed by a substantial increase by late 2000 (Fig. 3c and Fig. S2c). The spread of the strain from NCR to Mindanao (Region XI) can be traced back to the late 2004, while the spread from NCR to Visayas (Region VI) can be traced back to 2007 (Fig. 3c and Fig. S2c). Local spread in all three island groups increased by mid-2008 during the peak of *R_e_* (Fig. 3c and Fig. S2c). When *R_e_* rebounded during the late 2013 and onward, the strain was reintroduced to Luzon from Mindanao (Fig. 3c and Fig. S2c).

### CRF01_AE effective population size *N_e_* overtook that of subtype B around 2013, while the effective reproductive number *R_e_* of the two subtypes fluctuated out of phase

To contextualize the population dynamics of CRF01_AE, phylodynamic analysis was also performed on subtype B sequences available in the RITM Molecular Biology Laboratory database and by using the best-fitting relaxed clock model determined by the path sampling and stepping-stone procedure (Supplemental File XLSX), focusing on the largest monophyletic Philippine clade of subtype B sequences (Fig. S3). The resulting BSP analysis revealed an exponential growth phase in subtype B *N_e_* from 2003 until about 2010, during which it was comparable to the CRF01_AE *N_e_* (Fig. 4), followed by a peak and plateau phase wherein *N_e_* remained within the same order of magnitude from 2010 onwards (Fig. 4). The CRF01_AE *N_e_* had a longer lasting growth phase and a significantly higher peak than the subtype B *N_e_* by 2013 (Fig. 4). The estimated evolutionary rate of 2.524E-3 [95% HPD: 1.8829E-3, 3.1806E-3] substitutions/site/year and tMRCA of 1995.1435 [95% HPD: 1982.4503, 2003.6044] for subtype B were not significantly different from those of CRF01_AE (Table 1).

**Figure 4.**
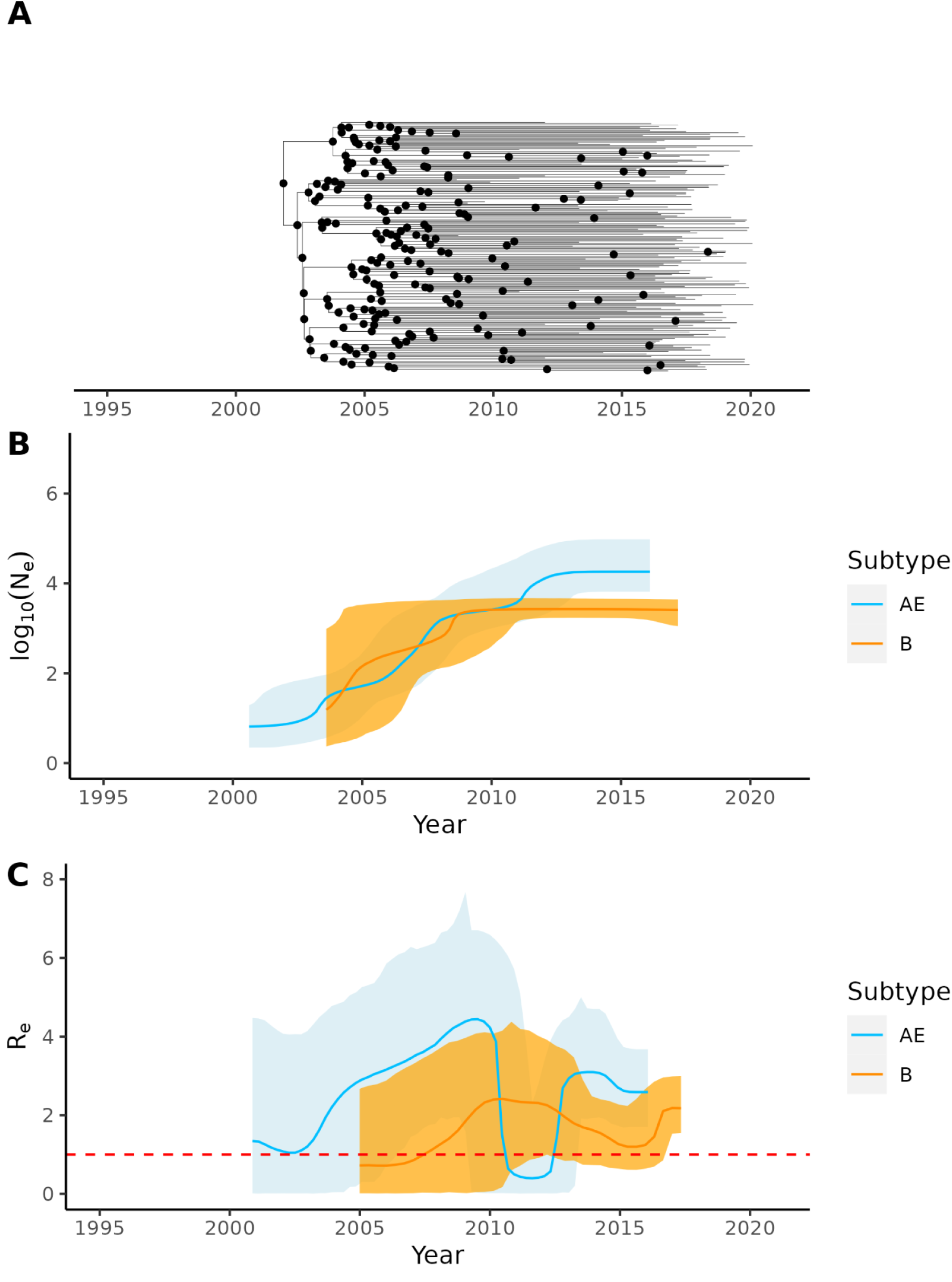
Comparison of phylodynamics between Philippine CRF01_AE and subtype B. (A) Time-scaled maximum clade credibility tree from BDSKY analysis of subtype B PR/RT sequences summarized with TreeAnnotator. Common ancestor nodes are highlighted as black dots. (B) The change in subtype B effective population size (*N_e_*) over time in log scale obtained from BSP analysis, superimposed over that of CRF01_AE, with the median represented as a black line and the 95% HPD as an orange-shaded interval. (C) The change in the effective reproductive number (*R_e_*) over time obtained from BDSKY analysis, superimposed over that of CRF01_AE, with the mean represented as a black line and the 95% HPD as an orange-shaded interval. A dashed red line indicates the value of *R_e_* equal to 1.

Phylodynamic analysis with the Birth–Death Skyline Serial model showed that the trend of the subtype B *R_e_* also fluctuated over time in a similar but lagging pattern compared with the *R_e_* of CRF01_AE (Fig. 2a; Supplemental File XLSX). During the first peak of the CRF01_AE *R_e_* around 2008 (3.71 [95% HPD: 1.71, 6.14] in 2008-04-10), the subtype B *R_e_* was still increasing (1.26 [95% HPD: 0.0225, 3.70] in 2008-01-28) (Fig. 2c). When the CRF01_AE *R_e_* transiently decreased around 2010–2011 (0.398 [95% HPD: 0.0105, 2.28] in 2011-07-16), the subtype B *R_e_* had reached its first peak phase (2.32 [95% HPD: 0.86, 4.15] in 2011-07-07) (Fig. 2c). When the CRF01_AE *R_e_* rebounded in 2013 (2.87 [95% HPD: 1.78, 4.22] in 2013-12-26), the subtype B *R_e_* was decreasing (1.63 [95% HPD: 0.73, 2.48] in 2013-11-30) until it increased again in early 2016 (2.10 [95% HPD: 0.85, 2.96] in 2016-04-25) (Fig. 2c). The peaks and troughs of the subtype B *R_e_* fluctuations were less in magnitude and steepness than those of CRF01_AE (Fig. 2c).

## Discussion

The HIV CRF01_AE phylogeny in this study revealed a large monophyletic Philippine clade, supporting a single past introductory event that led to the majority of infections in the country. This is consistent with the first-comer advantage and strong founder effect observed in HIV epidemics outside Africa [37]. Based on the phylogeny of sampled sequences, there is no evidence of much ongoing transmission from other sporadic introductions. The monophyly and estimated tMRCA of CRF01_AE are consistent with results from a study using nearly full-length genomes from the Philippines [15] and a recent reconstruction of CRF01_AE transmission from Africa to Asia [16]. The tMRCA estimates of the single largest CRF01_AE transmission cluster under different datasets and models were all close to each other at around the late 1990s to early 2000s, about a decade later than when the subtype expanded in Southeast Asia coming from Africa [16] and the earliest identified CRF01_AE sample in the country around the 1980s [10]. The estimated evolutionary rates for CRF01_AE from the BSP and BDSKY analyses were similarly robust at about 0.003 substitutions/site/year, very close to the estimate in a previous study when either the full coding region or the *gag* gene is used but not when the *pol* gene or nearly full-length genome is used [16], [37]. This could be due to different selected regions in the sequence or different epidemiological dynamics in the sampled locations [56].

In our analysis, the confidence intervals of the tMRCA and *R_e_* of the largest monophyletic CRF01_AE and B clusters overlap and thus are comparable or not significantly different from one another. However, the trend of the CRF01_AE *R_e_* was to always be above that of subtype B except during 2010–2012 when CRF01_AE *R_e_* decreased transiently. A recent study in 2022 by Salvaña et al. has shown evidence of significantly higher viral load in CRF01_AE infections compared to subtype B and to other circulating subtypes, which suggests potentially higher transmissibility of CRF01_AE infections and a mechanism for why CRF01_AE has become the predominant subtype in the Philippines instead of subtype B [9]. The trend of higher CRF01_AE *R_e_* in our study is consistent with the hypothesis of faster transmissibility over subtype B, although further analysis should be done to confirm these findings. Use of whole genome sequences instead of subgenomic sequences may lead to more precise estimates of these epidemiological parameters for each subtype [18] as well as allow for more accurate classification of subtypes and recombinants forms [17]. The same study by Salvaña et al also suggested higher rates of transmitted drug resistance (TDR) among CRF01_AE infections compared to other HIV subtypes, which if confirmed may be a mechanism by which the current antiretroviral treatment regimens in the country that were tailored for non-CRF01_AE epidemics selectively favor the survival and transmission of CRF01_AE [9]. Another explanation for the predominance of CRF01_AE may be due to its being the first to expand in the local sexually active MSM networks by chance, such as through first-comer advantage [57]. Both subtypes have been present in the Philippines since the 1980s, but earlier rapid growth of CRF01_AE over B was not observed in the country at that time [10]. Both of their largest transmission clusters were introduced prior to 2008 when most cases were from the heterosexual population [2], but our results suggest that a steep increase in CRF01_AE *R_e_* may have preceded that in B peaking around 2008–2009, shortly after which cases in the MSM population overtook those in the heterosexual population and during the rise in cases among 25–34 and 15–24 age groups relative to 35–49 year olds [2]. Consistent with being the first to expand in the susceptible sexually active MSM population, CRF01_AE was found in greater proportion (64%) than B (27%) in a subanalysis of MSM patients from infections between 2007 and 2012 [10], [11]. One might also speculate that the period of time wherein public health interventions targeted heterosexual/IDU rather than MSM risk groups early on in the epidemic may have favored more rapid expansion of the subtype that spread in the MSM population first if this occurred before policy change could adapt to the shift in the largest risk group. For example, in the 2009 country report to the UN Declaration of Commitment to HIV and AIDS, there was a 14% and 55% reach of prevention programs among the most-at-risk population in female sex workers (FSWs) in 2007 and 2009 compared to 19 and 29% in MSM, respectively [58]. From the same report, there was also a 65 and 65% proportion of condom use among FSWs in 2007 and 2009 compared to 32 and 32% in MSM, respectively [58]. In the HIV epidemic in China, the infection route of sexual contact was found to be more likely to be an infection with CRF01_AE compared to contaminated needles, showing that specific transmission routes and risk-groups can be dominated by a specific subtype [59]. Transmission in the MSM risk-group was also found to more likely form clusters [60] and has been linked to more occurrences of superspreaders than in non-MSM risk groups [61], [62]. Finally, it is also possible for a combination of multiple factors to have contributed to the predominance of CRF01_AE in the Philippines.

The CRF01_AE *R_e_* declined transiently in 2010–2012, overlapping with the 4th and 5th Philippine AIDS Medium Term Plans developed by the Philippine National AIDS Council for 2005–2010 [63] and 2011–2016 [58], respectively. Meanwhile, subtype B *R_e_* reached its first peak, which may have masked any more appreciable decline in cases in this period. Public health interventions have had a measurable effect on transmission based on modeling by the Philippine Epidemiology Bureau [1]. Perhaps this is what is reflected in the fluctuations in *R_e_,* in particular the shift in the focus of intervention to the MSM risk group during the 5th AMTP. Further evidence would be needed to confirm this association, and attribute the decline to specific interventions if not other contributing factors. It must be mentioned that drug resistance mutation genotyping at RITM was not performed from 2012 to 2013 due to lack of reagents, and neither were there CRF01_AE sequences from this time sourced from referring hospitals and social hygiene clinics. Thus, no CRF01_AE Philippine sequences from this period were included in this dataset for analysis. It is possible that undersampling of HIV sequences in this period could affect common ancestor nodes and parameter estimates in the years prior to it. However, the observed dip in CRF01_AE *R_e_* is likely to be genuine since decreases in the proportion of CRF01_AE cases relative to subtype B in 2010 and in the absolute counts of CRF01_AE cases in 2010–2012 were also observed in cohort study data from Telan et al. [11] and visualized by Salvaña et al. [10]. Additionally, both subtypes exhibited a decrease in *R_e_* in our analysis, and four (4) subtype B sequences sampled in 2012 belonging to the large subtype B transmission cluster were included in the phylodynamic analysis.

The peak and decrease in subtype B *R_e_* on the other hand lagged by a year, again consistent with cohort study data from Telan et al. [11] and visualized by Salvaña et al. [10] showing asynchronous peaks and declines of CRF01_AE and B infections over 2008–2013 [10]. One reason may be a later start of expansion of subtype B in the MSM population while remaining to make up a large if not dominant proportion in heterosexual/IDU cases [10], [11]. Perhaps interventions that caused a steep decline in the *R_e_* of the larger CRF01_AE transmission cluster among MSM had a delayed and more gradual effect on the smaller subtype B transmission cluster spreading in the same susceptible MSM population. It may also be of interest to investigate whether inter-subtype competition between CRF01_AE and B for susceptible hosts played a role in the opposed alternating pattern of their respective *R_e_*. A pattern of opposed and alternating oscillation was observed in the *R_e_* of competing Influenza A virus strains [64].

In 2013 onward, the CRF01_AE *N_e_* was significantly greater than that of B, which plateaued since 2009. Additionally, the CRF01_AE *R_e_* experienced a steep rebound in 2013, while that of B remained suppressed although not significantly different from each other given their wide confidence intervals. These agree with the study by Salvaña et al. [10], which showed a rebound and statistically significant shift in the predominant subtype to CRF01_AE by 2013. While further investigation on the cause of the rebound is needed, one suspect that could be considered from our phylogeographic analysis is inter-island reintroduction of CRF01_AE such as the Mindanao-to-Luzon transmission we identified around this time. Continued growth in Philippine island groups/regions outside Luzon/NCR could also be a factor, given diffuse transmission across locations. Local CRF01_AE spread in Visayas increased around this time while that in Luzon and Mindanao remained stable.

Still, other factors may explain the rebound, like behavioral changes facilitated by increased usage of mobile dating services [65]. It would be interesting to investigate whether this increased CRF01_AE *R_e_* and *N_e_* in 2013 was simultaneous with the date of increased usage of mobile dating services in the country. CRF01_AE *R_e_* remaining higher than that of B from 2013 to 2016 could have given CRF01_AE ample time to grow even more predominant in the MSM population, reaching 82% of cases since that time [66]. It is possible that the sequences in this dataset are less informative of *N_e_* closer to the latest sample collection date, or that increases in *R_e_* are not immediately reflected in *N_e_*. Thus, analysis with more recent sequences is needed to determine if the rebounded *R_e_* of subtypes CRF01_AE and B after 2013 and 2016, respectively, resulted in continued increases to their corresponding *N_e_* soon after.

For Luzon, particularly the NCR, to have been inferred the origin of the CRF01_AE epidemic is plausible since the most dense and urbanized cities in the Philippines are found therein. Citizens from various provinces regularly travel to the big cities such as Metro Manila for work [53], and the busiest and most highly connected airport in the country is in the NCR [67]. Following this was a complex pattern of diffusion between the three Philippine island groups (13 sampled Philippine regions). The relative rates of migration between locations could not be concluded to be different from each other, suggesting uniform import/export of CRF01_AE infections between island groups. This is also expected given the extensive interconnectedness and frequent travel of the Filipino population to and from different island groups/regions in the archipelago [47]. Such a pattern of CRF01_AE diffusion suggests the need for geographically wide coverage of control measures to suppress the overall HIV epidemic in the Philippines. As only limited samples were sequenced from Visayas and Mindanao relative to Luzon (Supplemental File XLSX), follow up analysis should be performed using a more uniform sampling of sequences from all island groups and Philippine regions over time to confirm these findings as well as achieve higher resolution phylogeography.

To summarize, we showed that the introduction of CRF01_AE into the Philippines was between the late 1990s and early 2000s, the majority of CRF01_AE sequences belong to a single cluster, the CRF01_AE viral population size *N*_e_ exceeded that of subtype B by 2013, the CRF01_AE *R_e_* peaked in 2008–2009 and in 2013 onward, the CRF01_AE *R_e_* transiently decreased from 2010 to 2012, the peaks and trough of subtype B *R_e_* lagged behind that of CRF01_AE, CRF01_AE spread diffusely from Luzon (NCR) to other Philippine island groups and regions, and CRF01_AE migration rates between island groups/regions are comparable with one another. The shift from subtype B to the more aggressive CRF01_AE, with its faster progression to advanced immunosuppression [9], [68] and its implications on treatment and control interventions, greatly highlights the need to be vigilant on changing phylodynamics of HIV subtypes in the Philippines. Similar analysis with a more updated dataset should be performed to elucidate how the events from the COVID-19 pandemic from 2020 to 2022 influenced the trajectories of the largest ongoing HIV-1 transmission clusters in the country. The results of our study characterize the CRF01_AE-predominant epidemic, providing context for the effectiveness of past and current public health measures and may inform future measures toward more effective control of the HIV epidemic in the Philippines.

## Data Availability

The datasets and the XML files used in this study can be found at https://github.com/mblbdmu/CRF01_AE-PH.

## Acknowledgements

The authors would like to acknowledge the help of the Epidemiology Bureau of the Philippine Department of Health for providing access to de-identified metadata from its national database that were relevant to this study. The authors would also like to thank Hasnat Sujon for providing valuable technical help in reviewing and improving earlier versions of the manuscript.

## Supplementary Data

Supplementary data contain the parameters used in running BEAST and BEAST2 analyses; list of HIV subtype CRF01_AE sequences sampled manually from phylogenetic tree to estimate CRF01_AE tMRCA; list of HIV subtype CRF01_AE sequences sampled uniformly across date and island group to reconstruct *N_e_*, *R_e_*, and phylogeography; list of HIV subtype B sequences available for analysis (no subsampling); distribution by island group and region of uniform subsample of CRF01_AE sequences; effective reproductive number of HIV subtype CRF01_AE and subtype B over time using Birth–Death Skyline analysis; and submission, sample, and accession IDs of subtype B and CRF01_AE sequences in the study.

## Supplementary Data

### Supplementary Figures

**Figure S1.**
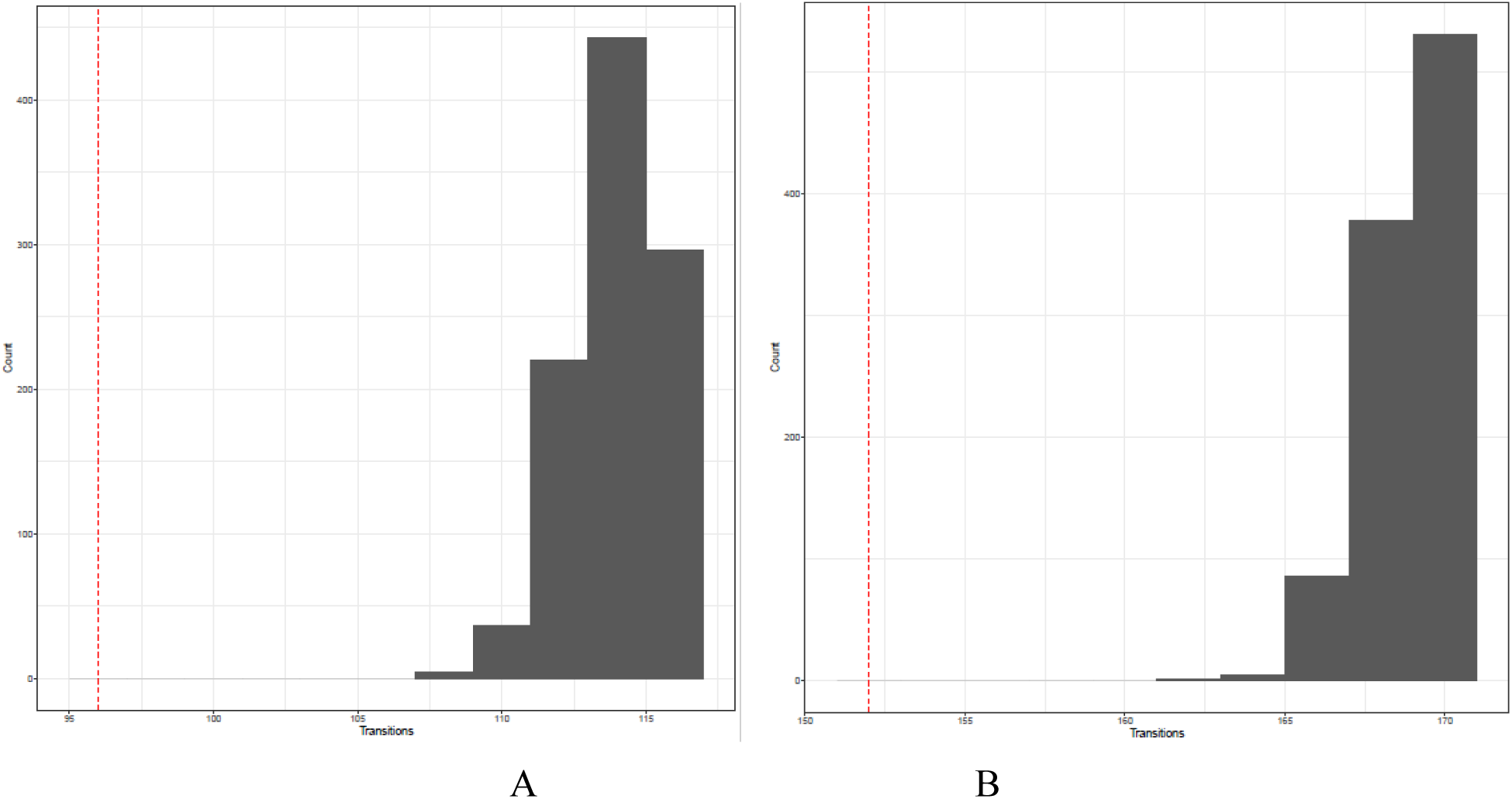
Slatkin–Maddison test for geographic clustering using the full set of Philippine CRF01_AE PR/RT sequences from the large CRF01_AE transmission cluster by (A) island group and (B) region. The dashed red line indicates the observed number of transitions. The gray histograms depict the distribution of transitions from the null model with 999 replicates.

**Figure S2.**
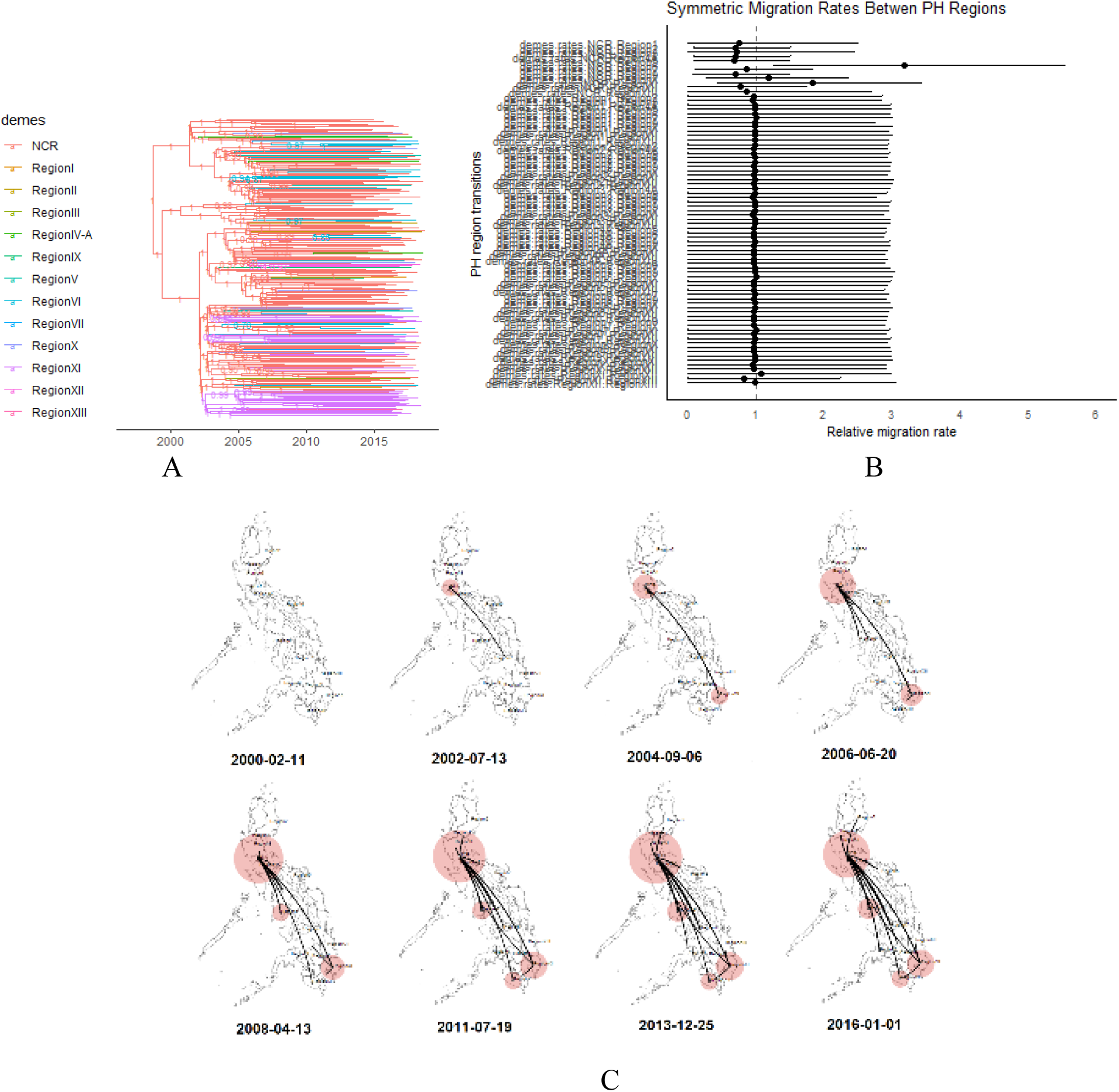
Analysis of geographic spread and relative migration rates of HIV CRF01_AE in the Philippines across PH administrative regions. (a) Maximum clade credibility tree of monophyletic PH CRF01_AE PR/RT sequences generated with BEAST1 under ‘phylogeo’ model and summarized with TreeAnnotator. Branches are labeled with posterior probability support values of corresponding nodes. Branches are colored according to the most likely geographic location of a branch at the level of PH administrative regions, wherein NCR (color red) was the most likely location of origin for the local epidemic followed by spread to other regions. (b) Forest plot of the relative migration rates of CRF01_AE between all pairs of PH administrative regions with a dashed line indicating a relative migration rate equal to 1.0. The 95% HPD intervals imply that there is no significant difference between the rate of migration of CRF01_AE between PH administrative regions. This suggests that no subset of migration rates dominate the diffusion process (i.e. no pair of locations that are the primary exporter-importer pair for CRF01_AE) and that there is extensive mixing between locations. (c) Phylogeographic spread of CRF01_AE between PH administrative regions over time visualized with SPREAD3. The size of the red polygons over the regions correspond to the intensity of localized virus transmission at the specified location and time.

**Figure S3.**
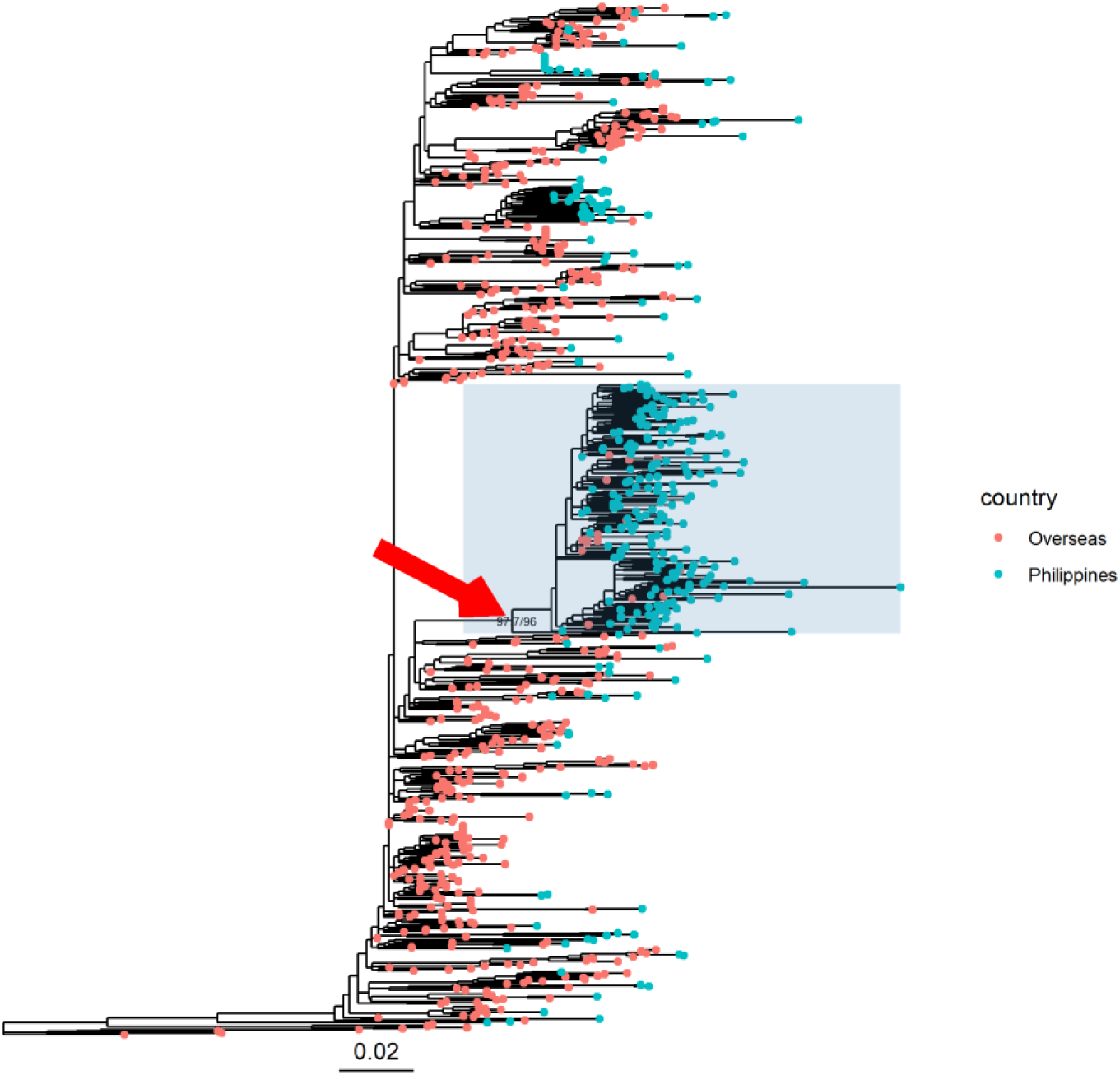
Maximum likelihood phylogeny of HIV subtype B generated with IQ-TREE2 using all eligible Philippine subtype B PR/RT sequences from the RITM MBL DRG database with a sampling period from 2008 to 2020, in the context of the closest subtype B sequences retrieved from LANL using HIV-BLAST and subtype C and A outgroup sequences. Tips of Philippines sequences from either the RITM MBL DRG or LANL databases are colored blue while overseas sequences from LANL are colored red. The red arrow indicates the node, with SH-aLRT and UFBoot branch support values, of the most recent common ancestor of the large monophyletic clade to which the majority of Philippine subtype B sequences belong. This clade is also emphasized with a blue rectangular highlight. The bottom scale bar shows a reference branch length for 0.02 substitutions/site. Overseas sequences contained within this cluster included those from Taiwan, Canada, United States, Japan, South Korea, Thailand, Australia.

### Supplementary Tables

**Table S1.**
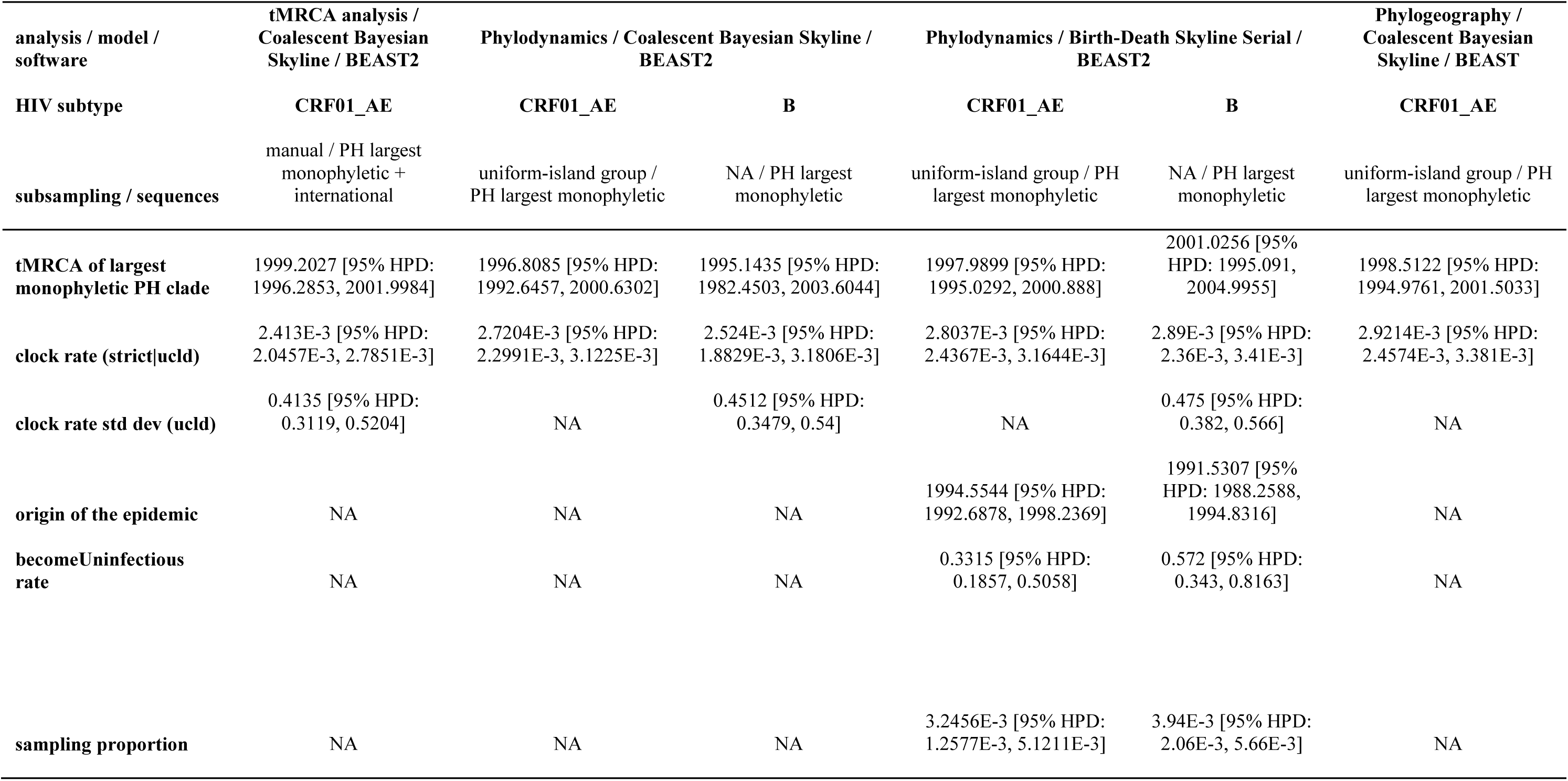
Posterior epidemiological and evolutionary parameter estimates from all BEAST and BEAST2 analyses performed in the study, including the mean and 95% HPD of each parameter under each analysis.

**Table S2.** BaTS analysis using island group as trait, 1000 trees downsampled from the CRF01_AE BSP sampled trees, and 999 simulated null trees.

| Statistic | observed.mean | lower.95..CI | upper.95..CI | null.mean | lower.95..CI.1 | upper.95..CI.1 | significance |
| --- | --- | --- | --- | --- | --- | --- | --- |
| AI | 9.14 | 7.82 | 10.45 | 14.00 | 12.86 | 15.08 | **<0.001** |
| PS | 54.35 | 50.00 | 58.00 | 72.14 | 69.33 | 74.42 | **<0.001** |
| MC (Luzon) | 13.39 | 11.00 | 17.00 | 7.92 | 6.44 | 10.29 | **0.014** |
| MC (Visayas) | 2.56 | 2.00 | 3.00 | 1.66 | 1.21 | 2.08 | **0.002** |
| MC (Mindanao) | 4.75 | 4.00 | 6.00 | 2.31 | 1.96 | 3.05 | **0.001** |

